## Supplementary Methods for "The clinical and molecular landscape of breast cancer in women of African and South Asian ancestry"

**Data collection:**

**Clinical data collation from Genomics England:**

For the clinical data for our cohort, the following tables within the Genomics England Research Environment were queried using LabKey via the R/LabKey API, using data release v17 (dated 30th March 2023). The relevant participant IDs were extracted from the *cancer_participant_disease* table through matching the cancer_disease_type "BREAST".

Other tables were queried using these unique Participant IDs as primary keys.

Primary data (from Genomics England):

Clinical:

- *participant*: basic participant data (year of birth, ethnic group, sex and gender)
- *cancer_staging_consolidated:* consolidated cancer staging information (T, N, M, diagnosis date, histology information, receptor status)
- *cancer_participant_tumour:* diagnosis date
- *cancer_invest_sample_pathology*: nodal status
- *cancer_registry*: cancer site and cancer event date
- *cancer_specific_pathology*: Allred scores for ER, PR, and HER2 status (including via fluorescence in-vitro hybridisation score for HER2)
- *death_details* and *mortality:* Date of death (if recorded)
- *av_imd*: Index of Multiple Deprivation (derived from GP postcode)
- *av_patient*: Survival status including date of death (if recorded)

Genomic:

- *cancer_analysis*: data regarding the germline and tumour samples and genomic analysis paths.
- *cancer_tier_and_domain_variants*: summarised somatic and germline variants as reported by Genomics England.

- *aggregate_gvcf_sample_stats*: information regarding the germline variant files; including the fraction of each determined ancestry in the vcf.

Secondary clinical data (NHS Digital and NCRAS):

- *hes_ae, hes_apc, hes_op*: Health Episode Statistics from NHS Digital: A&E, Admitted Patient Care and Outpatient Procedure information respectively
- *ecds:* Co-morbidity information from NCRAS.

One participant (out of the 3336 initial IDs) was in the database twice under two Participant IDs, flagged under a *duplicate_participant_id* entry in the table in both entries; one further participant withdrew consent before the v17 data release, leaving 3334 breast cancer participants within Genomics England.

*Collation of clinical data from tables in Genomics England:*

The collation of the clinical data from Genomics England proceeded in the following manner. For each of these categories, one single entry was consolidated for each participant, using the primary data source if available, and if not, the secondary was used, extracting the event or information from the record closest to diagnosis date, unless explicitly stated otherwise:

*Age at diagnosis*

- Primary data:
  - The diagnosis date from the *cancer_staging_consolidated* table was subtracted from the year of birth from the *participant* table
- Secondary data (used if primary was not available)
  - A diagnosis date was estimated from the secondary NHSD tables *hes_ae, hes_ap, hes_op,* where the visit was ICD-10 coded as C50* (malignant neoplasm of breast) or D05* (Carcinoma in situ of breast)
  - The estimated diagnosis date was subtracted from year of birth

*Self-reported ethnicity:*

Self-reported ethnicity was collated as a single ethnic group (White, Black or Black British, Asian or Asian British, Mixed, Other or Not Stated), and an ethnic subgroup within these groups using the NHS 2001 categorisation (https://www.datadictionary.nhs.uk/attributes/ethnic_category_code_2001.html).

- Primary data:
  - Ethnic categories taken from *participant* table

- Secondary data (used if primary data not available):
    - o Ethnicity taken as recorded from *hes_ae, hes_op and hes_apc* entries for each participant. If the ethnic groups agreed, then the collated ethnic group was taken to be the consensus; if not, the ethnic group was recorded as "Conflicted"; similarly for the ethnic subgroup.

*Tumour staging:*

Primary data:

- o Tumour staging (I, II, III, IV) taken from *cancer_staging_consolidated* table, along with tumour T, N and M staging, derived from imaging and pathology records for the participant.

*Tumour receptor status:*

- Primary data:
    - o ER, PR and Her2 statuses taken from *cancer_staging_consolidated*, derived from imaging and pathology reports.
- Secondary data (used if primary not available):
    - o ER, PR, Her2 Allred scores from *cancer_specific_pathology* were converted using the scale (0-2: Negative, 3 Borderline and 4-8: Positive). If Her2 score was Borderline, then the FISH score was used to derive Her2.

*Participant sex:*

- Primary data:
    - o Participant sex taken from *participant* table

*Survival information:*

- Primary data:
    - o Date of death taken from *death_details* table
- Secondary data (used for latest date of follow-up and for date of death if primary not available):
    - o Death and survival table taken from *mortality* and *av_patient* tables
    - o Hospital episode statistics in *hes_ae, hes_op and hes_apc* used to determine a date at last follow-up for survival analyses.

*Lymph node status and metastasis information:*

- Primary data:
    - o Site of metastasis taken from *cancer_participant_tumour_metastatic_site*

- Lymph node involvement taken from *cancer_invest_sample_pathology* pathology examination of lymph node biopsy

*Index of multiple deprivation:*

- Primary data:
  - Taken from *av_imd* table
- Secondary data (if primary not available):
  - Taken from NHS Digital secondary tables *hes_ae, hes_op* or *hes_apc*

*Primary trust/CCG and handling Genomic Medicine Centre information:*

- Primary data:
  - CCG ID, location and name taken from NCRAS secondary table *av_tumour*
  - GMC ID and name taken from *participant* table.

For the participant with duplicated IDs form two separate enrolments at different CCGs, the clinical information was checked to ensure that there were no discrepancies, aside from the CCG and GMC processing information and the date of diagnosis: the duplicate information was removed, and the earlier diagnosis date was taken, leaving a single row for this participant.

**Clinical data collation from Breast Cancer Now Biobank:**

321 participants in Genomics England were provided by the Breast Cancer Now Biobank and dually consented by both the BCN Biobank and Genomics England. For these participants, the clinical information held by BCN Biobank in their database (Date of birth, date of diagnosis, data of death – if present, ethnicity, receptor statuses) was collated for the entry containing the primary diagnosis of breast cancer and imported into the Research Environment. The clinical table we derived within the RE for these participants in these categories was updated with that from the BCN Biobank. The overlap between the BCN Biobank data and our analysis cohort was 195 participants. A comparison of age at diagnosis calculated as above using Genomics England data and that calculated solely using BCN Biobank data for the same participants, showed at most a discrepancy of one year between the two datasets, with an $R^2$ of 0.95.

This dually-consented subset of participants forms the basis of our BCN Biobank pharmacogenomic cohort.

**Genomic data collation from Genomics England:**

All genomic data was generated and provided by Genomics England through the trusted Research Environment. Samples of germline genetic material and tumour tissue were extracted, and Illumina sequenced, mapped to the hg38 genome assembly, and variants were called: germline variants via the Isaac single sample short variant caller and somatic short variants called using Strelka2 on the somatic and matched germline. Larger copy number changes and structural variants were called using the Manta (structural variant calling) and Canvas (copy number changes) pipelines. Genomics England provided reports of the small variants (SNVs and indels) called for both somatic and germline in the *cancer_tier_and_domain_variants* table, with locations for the raw data held in the *cancer_analysis* table. We used the small somatic variants from this table, but processed the raw germline vcfs directly using the Genomics England small variant workflow (available here: https://re-docs.genomicsengland.co.uk/small_variant/), v3.0.0. Briefly, this takes the raw germline vcfs, filters on the participants in the study and the relevant genes and annotates using the Ensembl VEP annotator (v107).

Finally, genetic ancestry (gAncestry) was called by Genomics England from the germline vcf for each participant, using a random forest classifier trained on the 20 principal components derived from 188382 good quality SNP locations within the 1000G phase 3 data. Each germline vcf was filtered for these locations, then projected onto the first 8 PCs, then 400 random trees were generated, each classifying the vcf into a gAncestry superpopulation (AFR: African, EUR: European, EAS: East Asian, SAS: South Asian, AMR: American Admix). The proportions of the 400 trees classifying the vcf into each superpopulation per participant were available from the *aggregate_gvcf_sample_stats* table, from which the gAncestry can be determined. If one superpopulation fraction was $\geq 0.8$, the participant's gAncestry was labelled as that superpopulation; if one was $\geq 0.5$ but $< 0.8$, the gAncestry was labelled as that superpopulation with an "_Admix" suffix; otherwise, the gAncestry was labelled as "Admix" (or undetermined).

Up-to-date information from EHRs, comprising structured and unstructured data, was accessible for the patients dually consented by both Genomics England and the BCN Biobank, with data from other resources showing propensity for being sparse and/or fragmented. A BCN Biobank pharmacogenomic cohort (n=160) was compiled from this dually-consented group, taken from those participants which were in the Genomics England EUR, AFR and SAS gAncestry cohorts.

**Genomic data collation from TCGA:**

Simple somatic variation data for all TCGA-BRCA patients was downloaded from the GDC data commons, and then filtered using the TCGA participant ID for our TCGA cohort. The gAncestry calls were accessed from the Carrot-Zhang study that defined the consensus ancestry call from five calling methods.

Germline data for the TCGA-BRCA cohort was downloaded from the PanCanAtlas from the analysis of 10.389 TCGA cancers and filtered for those participants which overlap our TCGA analyses cohorts. The germline data was also passed through a converted version of the Genomics England small variant workflow, filtering on the genes of interest and annotating using the Ensembl VEP annotator (v107).

**Genomic data collation from G&H:**

Germline data for the G&H cohort was available within its Trusted Research Environment for 44k participants (July 2021). This WES data release was filtered for our breast cancer cases and control participants.

**Variant calling comparison:**

The three genomic datasets used here use different methods for calling their variants. For germline short variants, the TCGA took consensus variants arising from multiple methods (MuSE, VarScan2, Pindel and HaplotypeCaller from GATK); Genomics England used the Isaac aligner-Starling single-sample variant caller pipeline for faster calling; and G&H used GATK HaplotypeCaller with hard filtering post-calling. All the variant callers used show similar sensitivities for SNVs and short indels on tissue samples), but TCGA's approach, taking a consensus call increases the confidence in their final released variants.

For the somatic small variants we use in this paper, TCGA again used a consensus method based on the results from MuSE, VarScan2, Pindel and Mutect2 from GATK, with Genomics England using the Strelka2 caller. Both Mutect2 and Strelka2 use a statistical model for calling variants from paired samples but differ in their application of the model: Mutect2 uses a Bayesian classifier and Strelka2 uses a trained machine learning method, but their sensitivities in calling variants on paired samples are very similar.

For the copy number changes and larger structural variants we use, Genomics England used Canvas for copy-number segmentation and Manta for structural rearrangements. The Canvas/Manta pipeline is very sensitive in determining variants of >200bp, but smaller changes (<200bp) are occasionally missed, particularly when comparing with SNP-based CNV methods such as ASCAT, which TCGA used for their CNV calls, which we have not used here.

**Data analysis:**

The analysis cohort was determined following the flow diagram of Figure 2. The criteria for exclusion are derived from the collated clinical and genomic data for the cohort as described in the previous section. The three problematic samples were identified at the QC stage.

*Demographic data from clinical tables*

To compare the demographic data (and germline BRCA status) from the AFR cohort and the SAS cohort against the EUR reference, the following method was used (in all cases gAncestry is an unordered factor with EUR as the reference level):

- For age at diagnosis and age at death, a linear model was tested:

$$Age \sim gAncestry$$

- For the other factors, the upper levels of the factor were compared against the reference level; because of the small numbers in AFR and SAS cohorts for lower levels, some factors were grouped in the following manner:
    - Tumour stage was split into stages I-II (reference level) and III+ (upper level)
    - Tumour grade was split into grades X (ungraded), 1 and 2 (reference level) and 3 (upper level)
    - IMD was split into quintiles 4-5 (reference level – least deprived), and 3, 2 and 1 (upper levels – most deprived quintiles)

  For receptor statuses, positive receptor status was taken as the reference level.

  For these factors, a logistic model was applied of the form:

$$Factor \sim gAncestry$$

  Where $Factor$ is 1 for the comparison level and 0 for the reference.

- For tumour mutational burden (once a value was calculated), a scaled negative binomial regression model of the following form was fitted:

$$Scaled\ TMB\ (\#\ mutations) \sim gAncestry$$

All analysis was performed using R.

To confirm the effect sizes in a direct AFR and SAS comparison, we separated these two groups, and re-ran the regressions on this reduced cohort.

*Tumour mutational burden calculation from genomic data*

Tumour mutational burden was calculated following the method in doi:10.1016/j.ccell.2022.08.022, correcting for gAncestry. The chosen gAncestry reference population within gnomAD59 was AFR for AFR, SAS for SAS and NFE (non-Finnish European) for EUR. Non-synonymous somatic variants within exonic regions were filtered based on the relevant gnomAD population frequency, the COSMIC database60 and the TOPMED (v3) database. A mutation was retained if it satisfied one of the following criteria: (i) gnomAD population frequency was ≤ 0.1% (rare) and the VAF was ≥ 3% or if the gnomAD population frequency ≤ 0.1% and the VAF was < 3% and annotated by at least two COSMIC identifiers or (ii) if the relevant gnomAD population frequency was > 0.1% but ≤ 10 and annotated by at least two COSMIC identifiers (Figure 8 of main text). The variant was discarded if it appeared in the TOPMED database at a frequency > 0.1%. The count of retained mutations per sample was divided by the size of the exome (≈ 35.4 Mb) and multiplied by $10^6$ to calculate the TMB per Mb of exome. It was at this stage that the three problematic samples were identified and removed, as inspection of the calculated TMB showed their resulting values were high outliers compared to the remainder of their superpopulation cohort.

*Germline variation determination (Genomics England cohort, TCGA and G&H)*

For Genomics England and TCGA, germline vcf files from our participants were passed through a pipeline to collate and annotate variants in our selected genes using bcftools and VEP (version 107). For our determination of pathogenicity, we restricted to the canonical transcript, and filtered further using the pathogenicity classification and confidence from ClinVar (Figure 9, main text).

For Genes & Health, the joint germline WES vcf, which had been previously annotated using VEP version 107, was filtered on the genes and the participants in our case and control and passed through the selection criteria in Figure 9. Because the Genes & Health cohort contained both male and female participants, the filtering and collation process prior to combining the individual WES vcfs failed on chromosome X, and so genes from our list located on chromosome X were not included in the Genes & Health case:control study.

As per the demographic calculation, a logistic model was fitted for the forest plots (with EUR in Genomics England/TCGA or control as the reference level for the $factor$):

$$variant \sim factor.$$

*Note that in the Genomics England cohort, the factor is the unordered gAncestry with EUR as the reference level.*

*Differential somatic variation determination*

Once the filtered variant list from the TMB calculation has been determined, the genes with variants were collated, and the individual variants were collated. For each variant gene and variant, a logistic regression was performed of the following form (comparing the three gAncestry groups jointly as an unordered factor):

$$Gene \text{ or } variant \sim gAncestry + Age\ at\ diagnosis$$

where the gAncestry reference level was EUR. If the adjusted p value for the $\beta$ coefficient relating to gAncestry was less than 0.1, the gene or variant was considered differentially present. However, in the cases where variants were not present in one of the two cohorts compared, the algorithm did not converge, so a second method was used. In this method, varied genes or variants were marked as present or absent in a cohort if present at above or below 2% in the respective cohort respectively (a threshold of 47/2343 in EUR, 3/138 in AFR and 3/123 in SAS). The three gene or variant lists present in each cohort above the threshold were intersected to give seven sets of intersections (EUR only, AFR only, SAS only, EUR+AFR (not SAS), EUR+SAS (not AFR), AFR+SAS (not EUR), all three cohorts). For our purposes, we took those genes or variants present at above 2% in AFR only and above 2% in SAS only, with the implication that they are present at <2% in the other two cohorts.

To control for the potential confounding factors of germline *BRCA1* and *BRCA2* status and IMD quintile in the logistic model, we added extra terms and restricted to those participants for which all the data was available:

$$Gene\ or\ variant \sim gAncestry + Age\ at\ diagnosis + BRCA\ status + IMD.$$

*Signature and HRDetect calculation*

We followed the method from https://github.com/Nik-Zainal-Group/signature.tools.lib (R library version 2.2.1): the somatic short-variant vcfs (locations of which are in the *aggregate_gvcf_sample_stats* table) were split into PASS SNVs and PASS indels without filtering; the somatic copy number vcfs were split into and CNVs. The R library was used to process the SNVs into SNV and DNV (dual nucleotide variant) catalogues, and the SVs into SV catalogues. Finally, the HRDetect function was applied on the four categories of somatic variants (SNPs, indels, SVs and CNVs) to compute the homologous recombination deficiency score using their pretrained classifier.

*Survival analyses (Genomics England cohort)*

Survival analysis was performed as in the recent pan-cancer Genomics England paper[51], using the code and methods provided, on our cohort, both as a whole, and split between the three gAncestry groups EUR, AFR and SAS.

**Code Availability:**

Although clinical and molecular data within the Genomics England TRE have been anonymised, the data are sufficiently detailed that the data could be deanonymised through data linkage, and thus cannot be extracted directly from the TRE. Codes for the analyses from this manuscript are available from the authors on request, but will need importing into the TREs for approved Genomics England Research Network and Genes & Health Research Network participants.
