## Supplementary Figures for "The clinical and molecular landscape of breast cancer in women of African and South Asian ancestry"

#### a. Genomics England

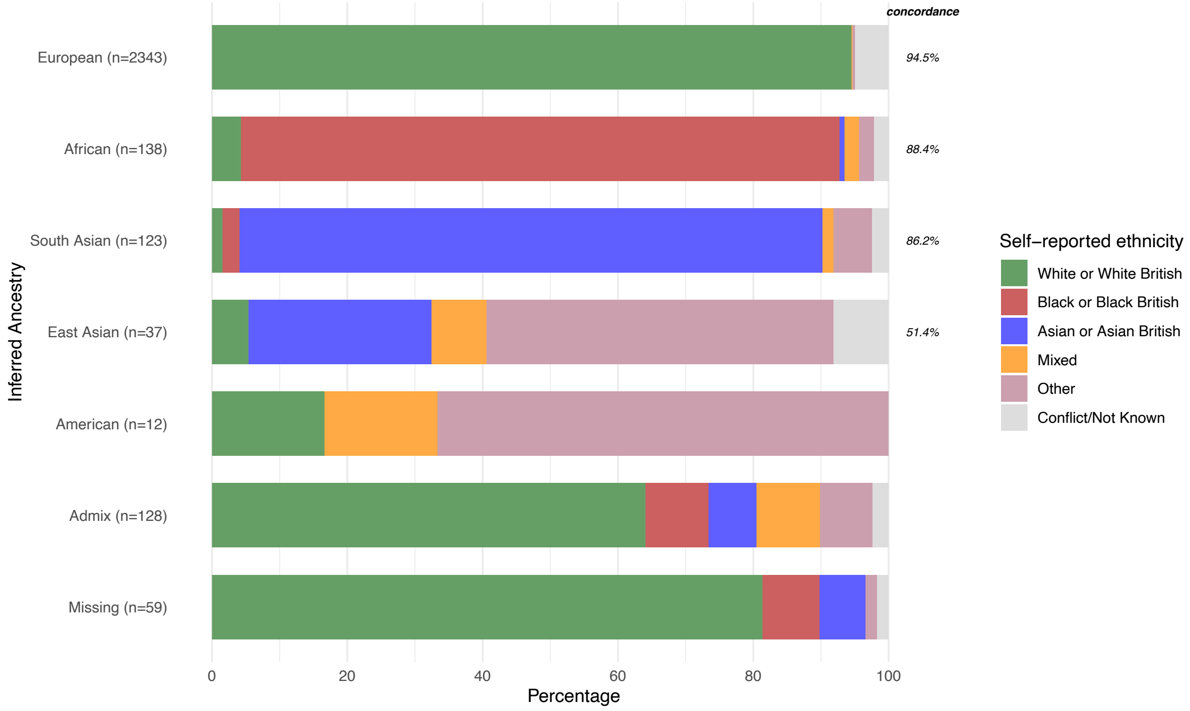


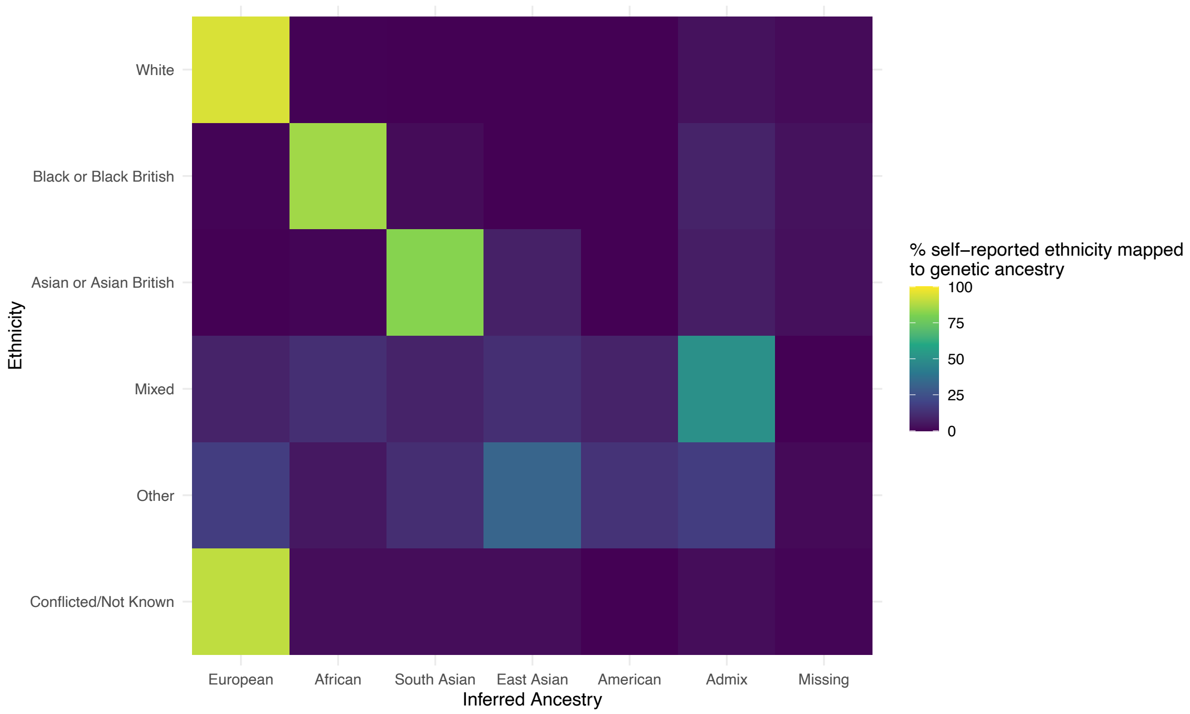

#### b. TCGA

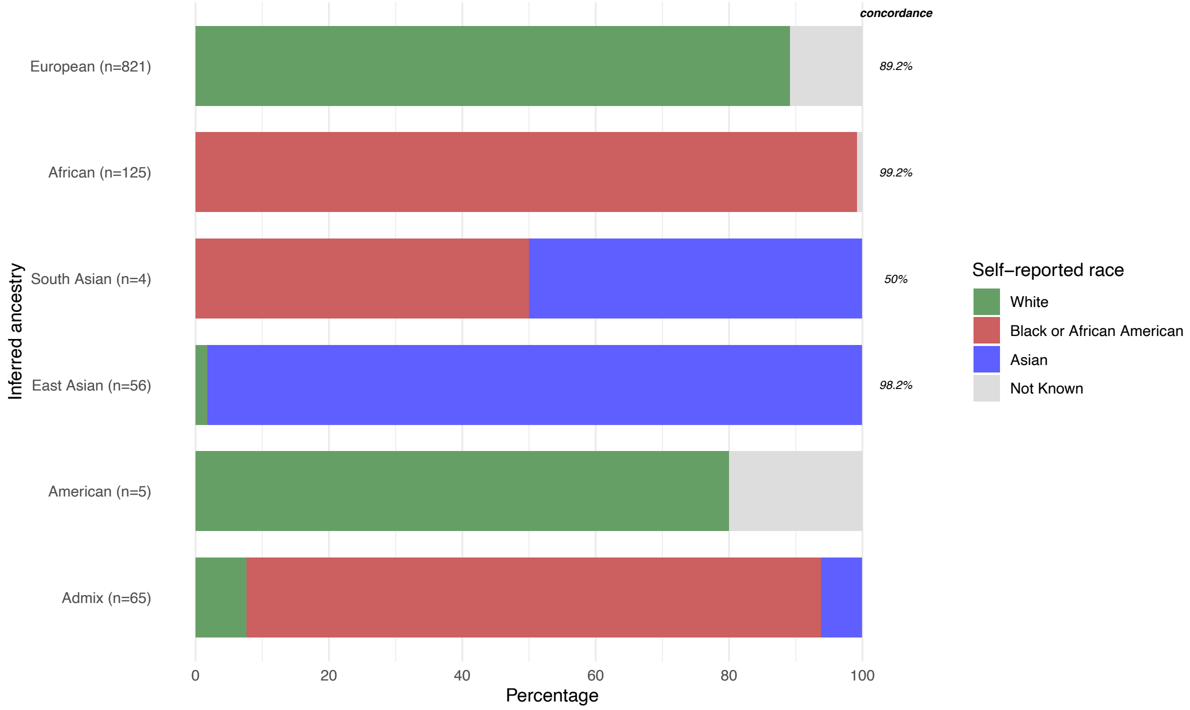
concordance
European (n=821)
89.2%
African (n=125)
99.2%
South Asian (n=4)
50%
East Asian (n=56)
98.2%
American (n=5)
Admix (n=65)
Inferred ancestry
0
20
40
60
80
100
Percentage
Self−reported race
White
Black or African American
Asian
Not Known

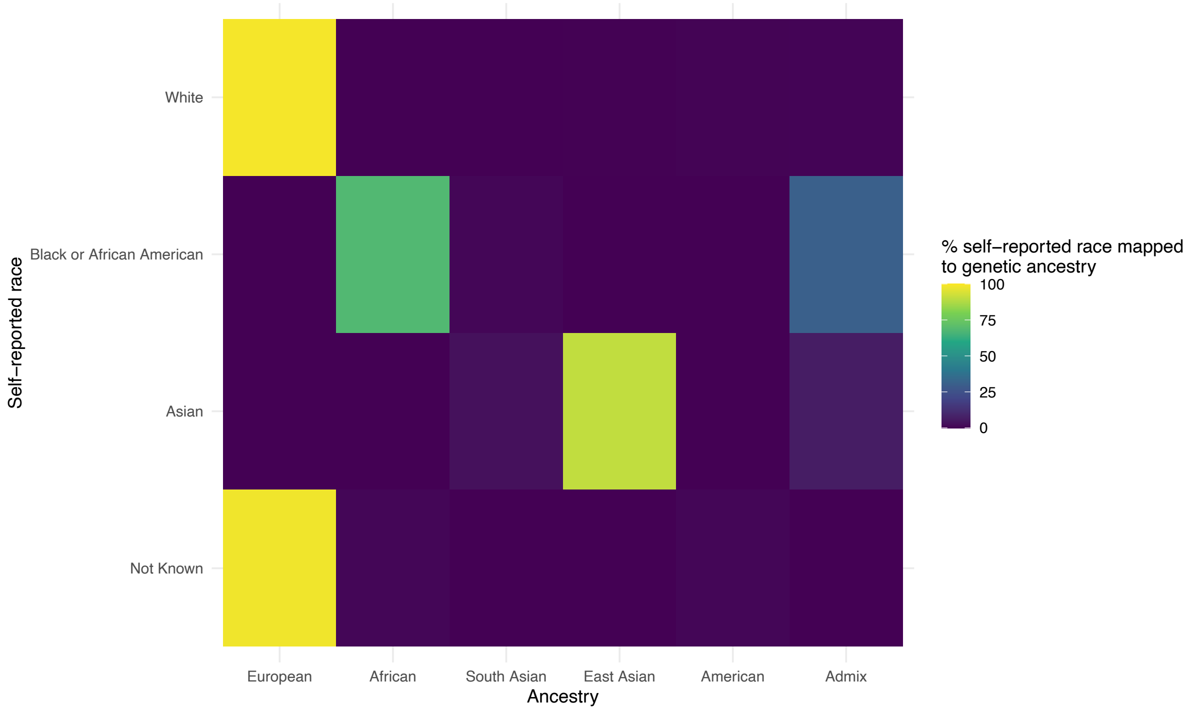
White
Black or African American
Asian
Not Known
Self−reported race
European
African
South Asian
East Asian
American
Admix
Ancestry
% self−reported race mapped to genetic ancestry
100
75
50
25
0

#### c. Genes & Health

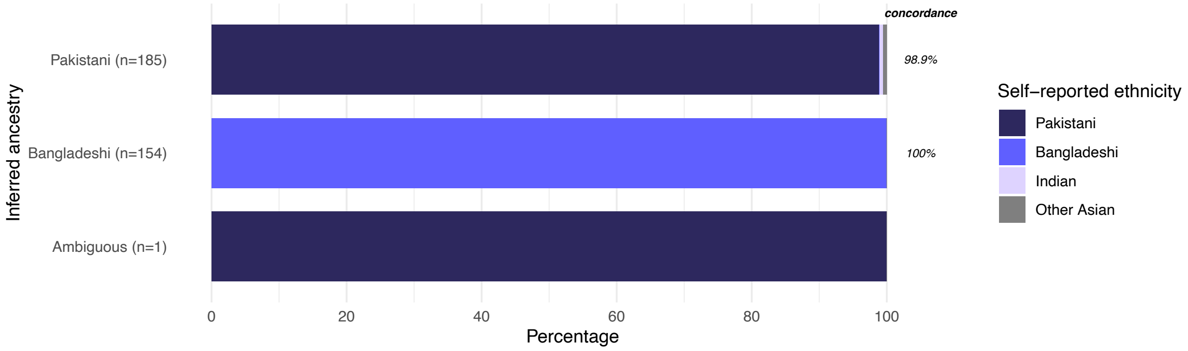


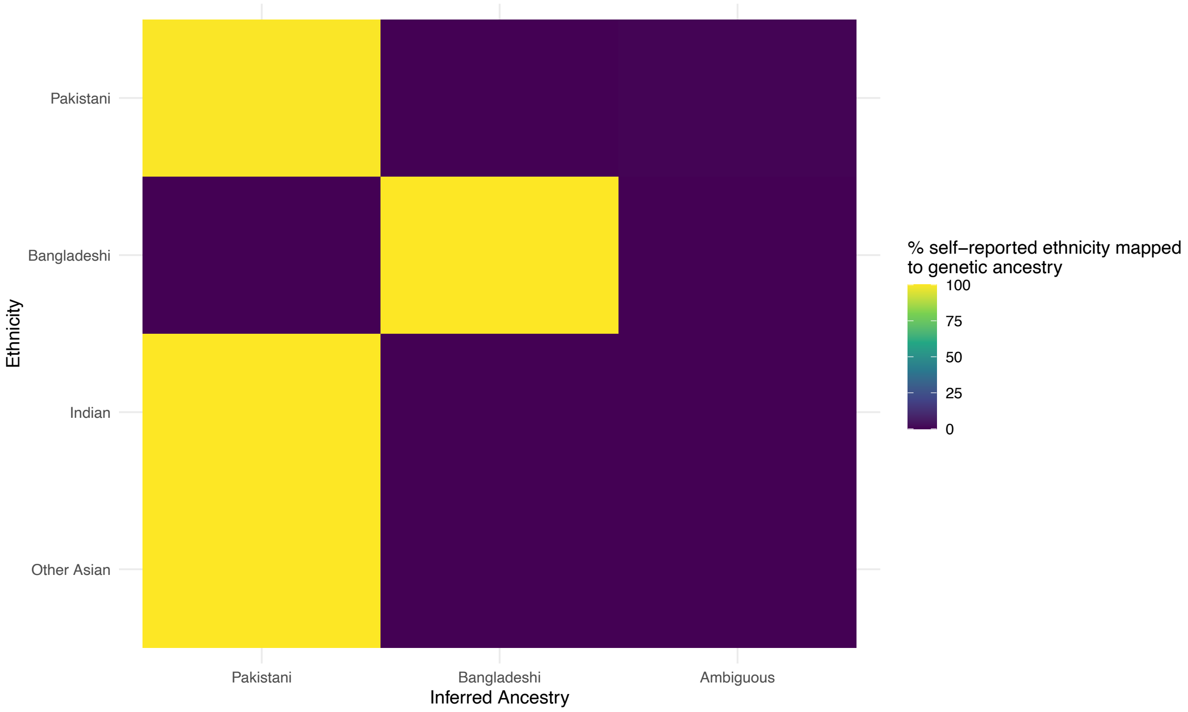


**Supplementary Figure 1.** Concordance between self-reported ethnicity/race and inferred gAncestry: **a.** in the Genomics England cohort; **b.** in the TCGA breast cancer cohort; **c.** in the G&H breast cohort at the subpopulation level.

**a.**

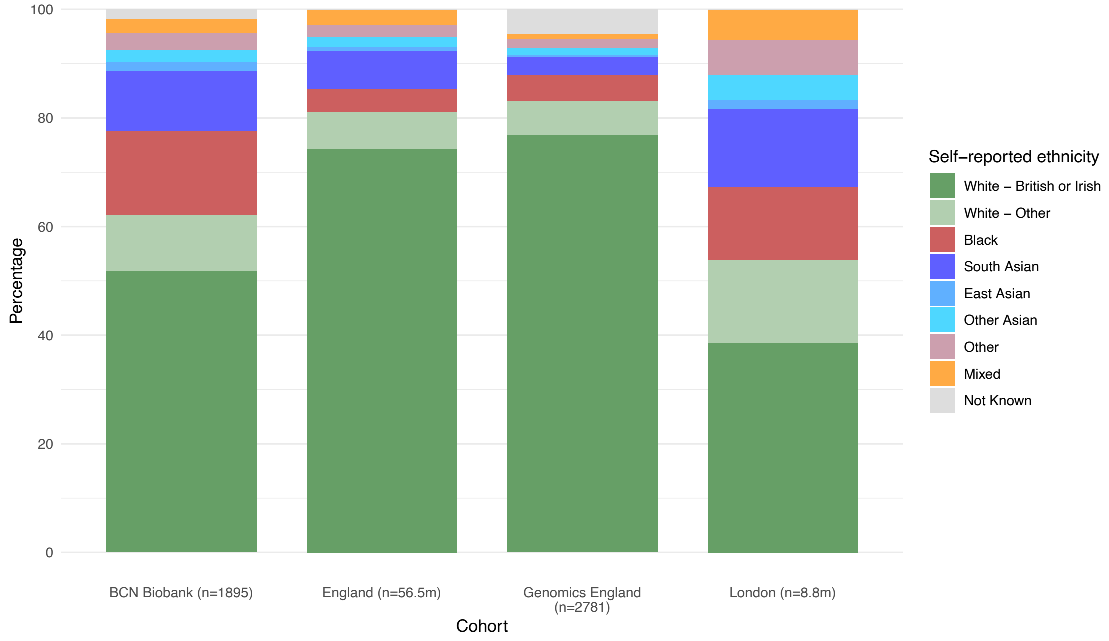


**b.**

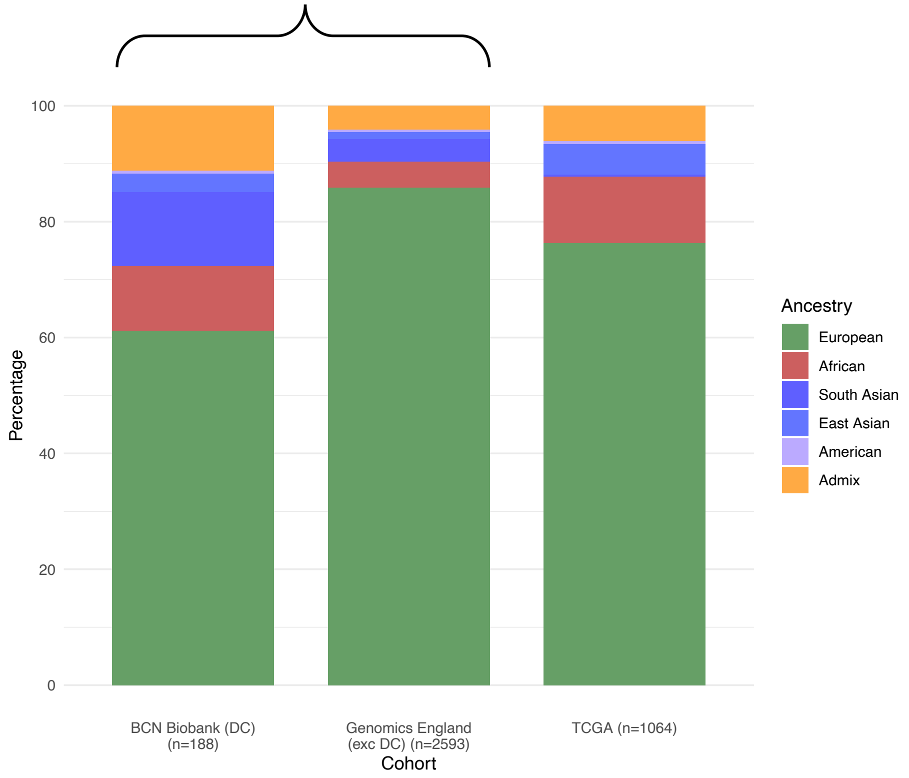


**Supplementary Figure 2.** Ethnic and gAncestry compositions of analysis cohorts. **a.** Ethnic composition of England, London (from 2021 UK Census), Genomics England analysis cohort and BCN Biobank validation cohort. **b.** gAncestry composition of Genomics England analysis cohort (excluding BCN Biobank dually-consented cohort), BCN Biobank dually-consented cohort and TCGA validation cohort

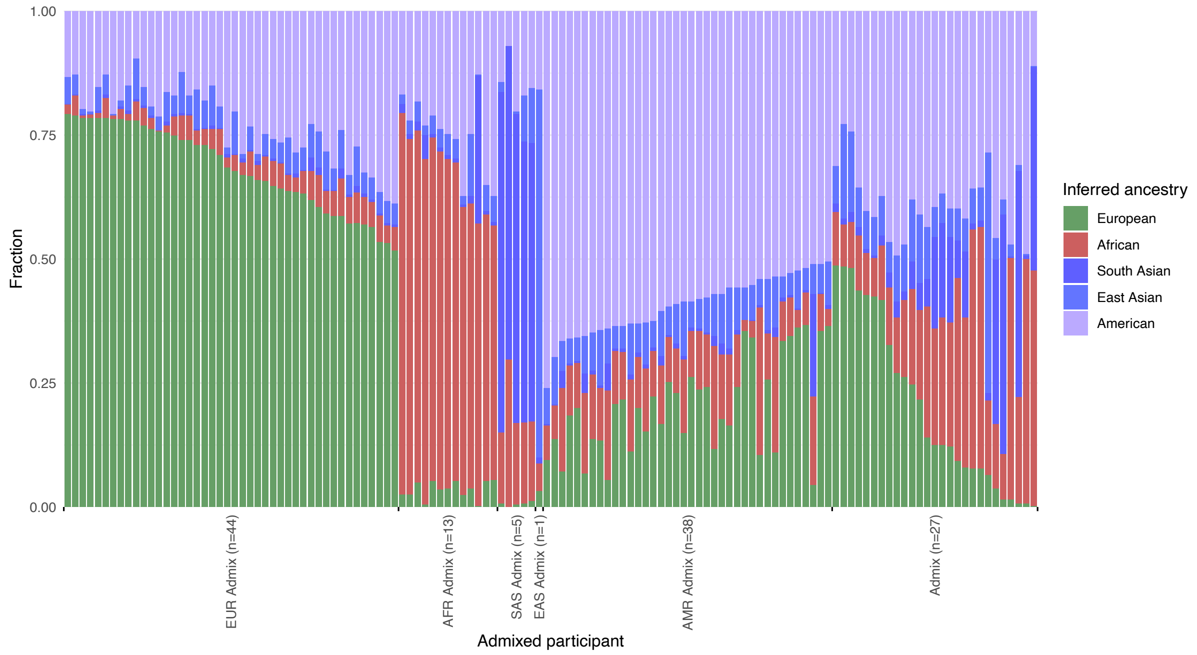


**Supplementary Figure 3.** gAncestry fractions per admixed participant within the Genomics England breast cancer cohort.

### a. TCGA

#### AFR v EUR

| Age | # EUR/# AFR | |
|---|---|---|
| Age at diagnosis | 812 / 122 | -4.08 (-6.58, -1.58) |
| **Age group** | | |
| 50 or over v Under 50 | [611/201] / [81/41] | 0.65 (0.432, 0.977) |
| **Menopause status** | | |
| Peri v Pre | [23/165] / [3/28] | 0.769 (0.216, 2.73) |
| Post v Pre | [553/165] / [67/28] | 0.714 (0.444, 1.15) |
| **Survival status** | | |
| dead v alive | [117/695] / [15/107] | 0.833 (0.469, 1.48) |
| **Stage** | | |
| II v I | [443/144] / [76/21] | 1.18 (0.7, 1.98) |
| III v I | [191/144] / [21/21] | 0.754 (0.397, 1.43) |
| IV v I | [15/144] / [3/21] | 1.37 (0.366, 5.14) |
| **TNM (T)** | | |
| T2 v T1 | [457/217] / [73/30] | 1.16 (0.733, 1.82) |
| T3 v T1 | [107/217] / [15/30] | 1.01 (0.523, 1.97) |
| T4 v T1 | [30/217] / [4/30] | 0.964 (0.318, 2.93) |
| **TNM (N)** | | |
| N1 v N0 | [262/383] / [35/67] | 0.764 (0.493, 1.18) |
| N2 v N0 | [91/383] / [11/67] | 0.691 (0.351, 1.36) |
| N3 v N0 | [62/383] / [5/67] | 0.461 (0.179, 1.19) |
| **TNM (M)** | | |
| M1 v M0 | [17/700] / [3/77] | 1.6 (0.46, 5.6) |
| **Receptors** | **-/+** | |
| ER | [143/626] / [47/71] | 2.9 (1.92, 4.37) |
| PR | [222/544] / [58/60] | 2.37 (1.6, 3.51) |
| Her2 | [440/124] / [51/11] | 1.31 (0.661, 2.58) |

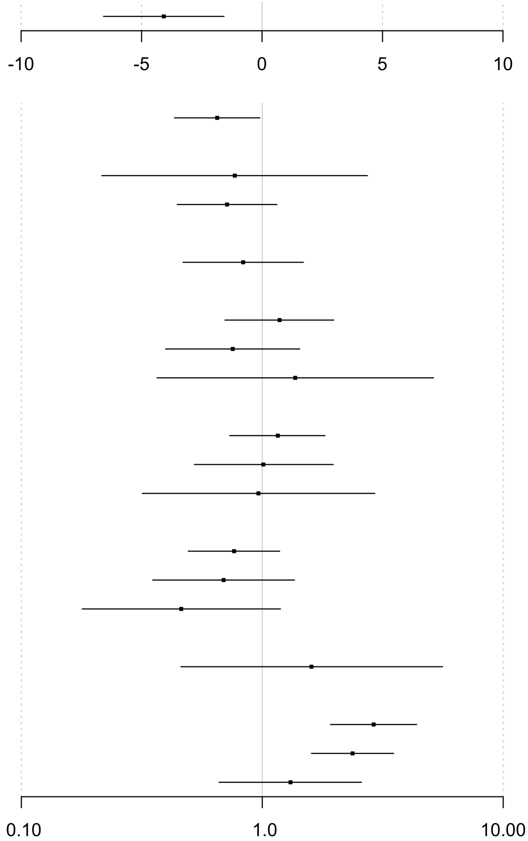


### b. BCN Biobank

#### Black or Black British v White

| Age | # White/# Black | |
|---|---|---|
| Age at diagnosis | 1176 / 293 | -2.54 (-4.15, -0.932) |
| Age at death | 192 / 59 | -6.48 (-10.6, -2.39) |
| **Age group** | | |
| 50 or over v Under 50 | [924/252] / [204/89] | 0.625 (0.47, 0.832) |
| **Menopause status** | | |
| Peri v Pre | [74/222] / [24/71] | 1.01 (0.596, 1.73) |
| Post v Pre | [733/222] / [158/71] | 0.674 (0.491, 0.926) |
| **Grade** | | |
| G2 v G1 | [172/30] / [30/4] | 1.31 (0.43, 3.98) |
| G3 v G1 | [136/30] / [45/4] | 2.48 (0.829, 7.43) |
| **Nodal status** | | |
| Positive v Negative | [307/645] / [109/130] | 1.76 (1.32, 2.35) |
| **Receptors** | **-/+** | |
| ER | [127/866] / [43/183] | 1.6 (1.09, 2.34) |
| PR | [98/336] / [25/70] | 1.22 (0.736, 2.04) |
| Her2 | [839/136] / [183/44] | 0.674 (0.463, 0.982) |

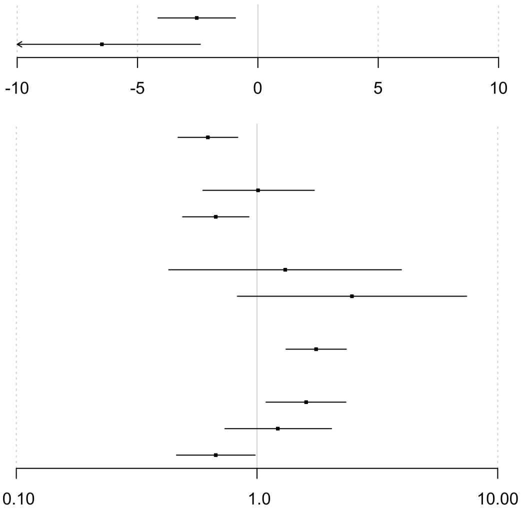

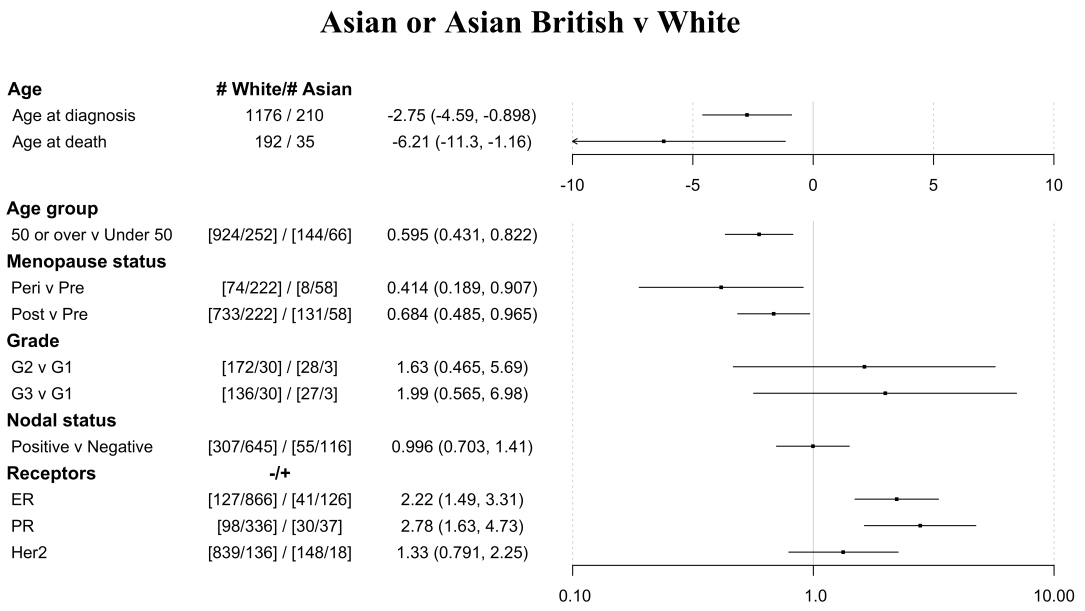


**Supplementary Figure 4.** Odds ratios of the clinical and molecular features of **a.** AFR patients of the TCGA against EUR patients and **b.** Black/Black British and Asian/Asian British patients in the BCN Biobank that have not been dually consented with Genomics England against White patients.

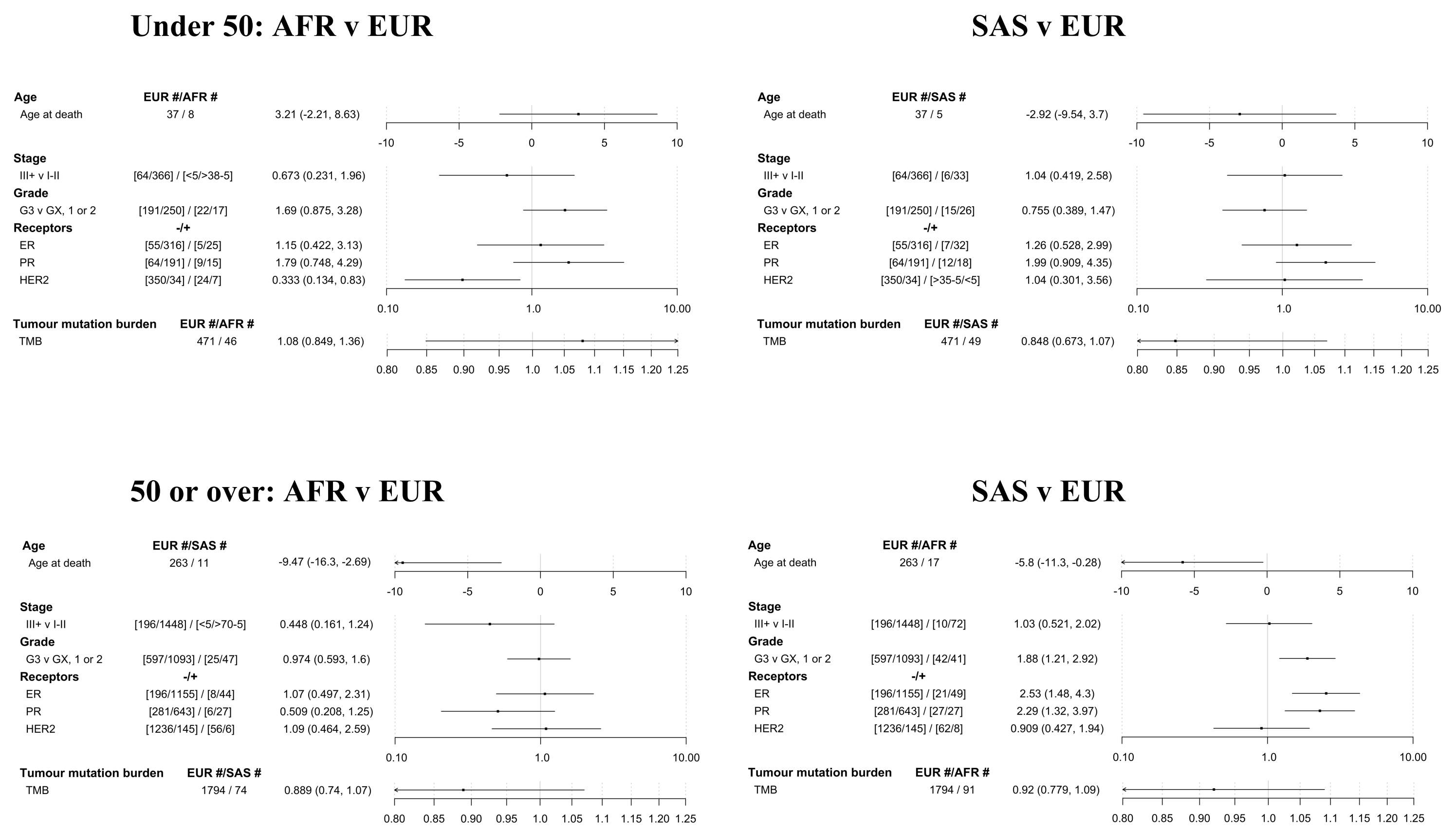


**b.**

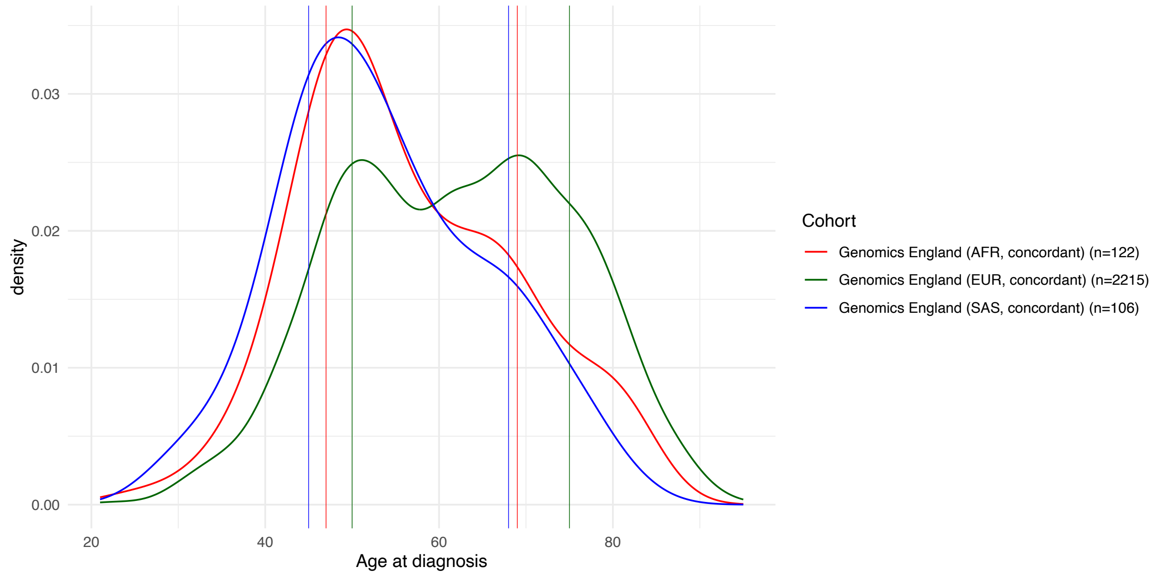


**Supplementary Figure 5. a.** Forest plots of the clinical and molecular features of non-EUR cohorts split based on a 50-year-old cut-off. EUR cohort split 471/1794 (under 50/50 and over); AFR cohort split 46/91 and SAS cohort split 49/74 respectively. **b.** Age distribution for patients in Genomics England with concordant ethnic group and gAncestry assignments. Application of the gAncestry-derived screening windows to the ethnicity-stratified cohort shows that the age groups proposed continue to encompass the central 60% of the age distributions intervals (AFR 47-69 years – 60.16%, EUR 50-75 years – 59.40%, SAS 45-68 years – 60.38%).

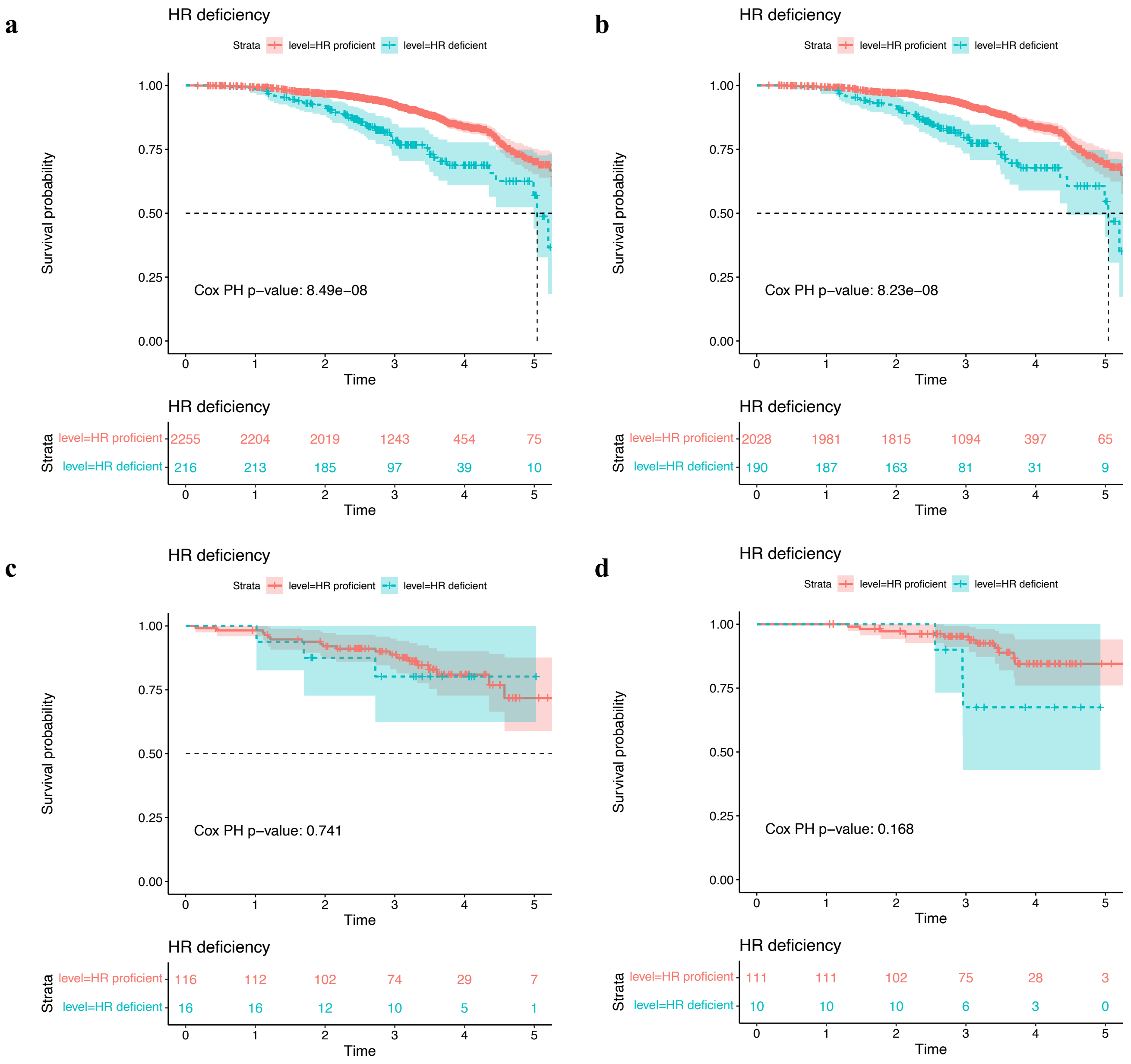


**Supplementary Figure 6.** Survival analyses of cohort based on HRDetect determination of homologous recombination deficiency: **a.** Whole analytic cohort with survival data (n=2471); **b.** EUR patients only (n=2218); **c.** AFR patients only (n=132); **d.** SAS patients only (n=121).
